# Hypertension Self-Management and Stroke Recovery Among Rural Adults in the Stroke Belt: A Mixed-Methods Study

**DOI:** 10.1101/2025.06.12.25329283

**Authors:** Mudasir Andrabi, Betty Key, RN Chaitali Dagali, Kayla Glass, Stephine Hart, Susan Appel, Lin Chen, Karlene Ball

## Abstract

Limited knowledge exist on HTN-related knowledge and health behaviors among African American adult stroke survivors with hypertension condition living in rural Alabama. To address this gap, we conducted a small pilot study with a mixed methods design for needs assessment of stroke survivors with a hypertension condition living in rural areas of Alabama. We followed the community engagement strategy approach to conduct our study.

After the approval from the Institutional Review Board, participants were recruited (N=25) using convenience sampling. We conducted surveys, followed by sequential interviews of our participants. Our needs assessment focused on knowledge and actual behaviors related to hypertension management among this population.

This paper presents findings from the quantitative and qualitative data collected for this needs assessment study. Data collection included: (i) HTN Knowledge-Level Scale test (HK-LS), (ii) HTN Self-Care Activity Level Effects (H-SCALE), and (iii) The Southampton Stroke Self-Management Questionnaire. Interested participants ( n=14) were interviewed using a PI-developed semi-structured interview guide.

Descriptive and inferential statistics were used to analyze the data collected from the surveys. The majority of participants (76%) had low level knowledge related to hypertension. Most participants also demonstrated limited adherence to hypertension behaviors including hypertension medication adherence (76%), DASH Diet (84%), physical activity ( 56%), and 72% had smoking habits. Our findings from qualitative interview data revealed the major themes of lack of knowledge related to hypertension management and post-stroke life management, lack of adherence to prescribe hypertension treatment, lack of continuity of care after discharge from hospital, and lack of social support.

These results indicated poor adherence to prescribed hypertension management behaviors. These findings highlight the need for a larger-scale study to assess heart health knowledge further and to identify the specific needs and preferences of this underserved population, an essential step toward developing tailored, community-informed interventions.

## Introduction

Stroke is a leading cause of long-term disability in the United States, with approximately 7 million Americans living with the debilitating effects of stroke.^1,2^ Stroke profoundly impacts physical, cognitive, and emotional well-being, with survivors facing significant challenges in recovery and reintegration into their communities. These challenges are especially difficult for individuals in rural and underserved areas, where barriers extend beyond healthcare access to include socioeconomic burdens, limited rehabilitation resources, and inadequate community-based support systems.^3-71^

The “Stroke Belt” refers to a multi-state region in the southeastern US where stroke mortality rates exceed the national average.^2^ The intersection of race and rurality further compounds these disparities, as the Stroke Belt has higher proportions of non-Hispanic African American and rural residents compared to other regions. Rural communities face higher stroke incidence, poorer risk factor control, and greater obstacles to preventive care and timely treatment.^7^ Geographic isolation, healthcare workforce shortages, and socioeconomic disadvantages further exacerbate stroke-related morbidity and mortality.^8^

African American (AA) adults bear a disproportionate burden of stroke, with incidence rates nearly double that of non-Hispanic White adults along with higher rates of stroke mortality.^3^ Hypertension (HTN), a primary modifiable risk factor for both incident and recurrent stroke, is more prevalent among AA adults, yet control of HTN is generally poor among stroke survivors.^6^ Effective HTN control is critical for secondary stroke prevention, but rural AA stroke survivors living in the Stroke Belt face compounded social and structural barriers that hinder timely care access, chronic disease self-management, and post-stroke recovery. Addressing these disparities requires a deeper understanding of the social, environmental, and healthcare system factors shaping survivors’ health trajectories.

Community engagement in stroke prevention offers a promising avenue for addressing these disparities. By leveraging social networks, local resources, and inclusive environments, community-driven stroke prevention efforts can improve long-term outcomes.^4^ However, research in this area remains limited. Existing studies primarily focus on stroke survivors’ lived experiences without capturing measurable patterns in self-management behaviors, healthcare utilization, and HTN control, key factors in developing tailored interventions.

To address these gaps, this study employed a mixed-methods design to provide a more comprehensive understanding of AA stroke survivors’ experiences and HTN self-management knowledge and practices. Specifically, this study aimed to 1) assess stroke survivors’ knowledge and adherence to hypertension self-management practices; and 2) explore survivors’ experiences with the healthcare system and perceived barriers to care. Findings from this study provide critical insights into the real-world challenges survivors face and serve as a foundation for developing future community-engaged interventions.

## Methods

### Design

This study employed a cross-sectional, descriptive, mixed-method design. Before its initiation, approval was obtained from the university’s Institutional Review Board (IRB# 22-09-5982). A descriptive analysis of the quantitative data collected from the surveys, and a thematic analysis of the qualitative data was conducted to explore participants’ experiences with healthcare services received during and after their stroke events. Findings from both methods were triangulated to enhance the depth of understanding and ensure a more comprehensive interpretation of the data.

### Participants and Setting

African American (AA) stroke survivors from three counties in Alabama (Hale, Sumter, and Pickens) were invited to participate in the study. These counties are designated as rural by the U.S. Census Bureau and classified as Health Professional Shortage Areas.^5^ Participants were recruited through community-engaged research practices. While we acknowledge that outreach activities represent an early point on the community engagement continuum, our work was intentionally designed in partnership with community stakeholders. These efforts went beyond outreach by incorporating community input into design, recruitment, implementation, and dissemination, and they serve as a foundation for sustained collaboration. Rather than being merely community-based, this study reflects a strategic CE approach that prioritizes relationship-building and positions the community as a partner in the research process.

We advertise our study using flyers and word-of-mouth, with a focus on individuals living independently in the community. Those residing in assisted living, nursing, or rehabilitation facilities were excluded. To facilitate recruitment, trusted community members living within these counties helped identify potential participants and connected study investigators to them at accessible community centers for eligibility screening. After explaining the study objectives and procedures, informed consent was obtained from each eligible participant.

#### Inclusion criteria

Participants were eligible for the study if they: (a) self-identified as being AA, (b) were age ≥ 50 years, (c) resided in one of three rural counties in Alabama during and after their stroke (Hale, Sumter, Pickens), (d) diagnosed with any type of stroke by a licensed healthcare provider (e.g., physician, nurse practitioner), (e) having a diagnosis of HTN and taking at least one prescribed antihypertensive medication, (f) able to communicate in English, (g) and demonstrating intact cognitive ability (a score of 22 or above on the Mini-Mental Status Examination scale^11^).

#### Exclusion criteria

Exclusion criteria were: (a) cognitive impairment as assessed by the Mini-Mental Status Examination ^6^ and (b) any suicidal ideation identified through the Beck Depression Inventory.^7^ A total of N=25 participants who met the eligibility criteria provided informed consent and were enrolled in the study. All participants (N = 25) completed quantitative surveys administered via Qualtrics. Following survey completion, a subset of participants (n = 14) also took part in sequential qualitative telephone interviews.

### Data Collection

Using four quantitative surveys, data was collected regarding participants’ socio-demographic information, their knowledge of HTN, adherence to HTN management self-care activities, and post-stroke self-management competencies.

#### Surveys

1. *Demographic survey:* An investigator-developed survey was used to collect socio-demographic information, including age, years of education, annual income, marital status, number of children, living situation including living alone ( yes, or no) , health insurance status (yes, no)), self-rated health ( good to excellent, fair to poor), number of years living with a diagnosis of HTN and stroke. Participants also indicated their county of residence. All data were self-reported and categorized for analysis.
2. *HTN Knowledge-Level Scale test (HK-LS)*: The HK-LS is a well-established measure designed to assess general knowledge of HTN, medical treatment and compliance, lifestyle factors affecting blood pressure (BP), diet, and HTN-related complications. The scale has high validity, with a Cronbach’s alpha of 0.85 indicating strong internal consistency, construct validity, and stability over time. The scale consists of 22 items across six sub-dimensions: “definition” (2 items), “medical treatment” (2 items), “drug compliance” (4 items), “lifestyle” (5 items), “diet” (2 items), and “complications” (5 items). Each correct answer earned 1 point, yielding a total score range of 0 to 22. ^8^
3. *HTN Self-Care Activity Level Effects (H-SCALE)*: The H-SCALE measures daily adherence to self-care activities for managing HTN, with strong internal consistency ( Cronbach’s alpha between 0.71 - 0.87). It assesses several self-care behaviors: medication adherence (3 items, score range 0-21), body weight management (10 items), adherence to the Dietary Approaches to Stop Hypertension (DASH) diet (11 items, score range 0-77), regular physical activity (2 items, score range 0-14), smoking (2 items, score 0-14), and abstinence from alcohol intake (3 items, score 0-14).^9^
4. *The Southampton Stroke Self-Management Questionnaire (SSSMQ):* The 28-item SSSMQ^15^ was used to measure participants’ self-management competencies regarding stroke. The scale has high internal consistency (Mokken r = 0.89) and excellent test-retest reliability (ICC = 0.928). Each item is rated on a 6-point Likert scale (1 = always false, 6 = always true) to measure attitudes, skills, and behaviors necessary for managing health and well-being after stroke.^10^

Qualitative interviews (45-60 minutes) were conducted using a semi-structured questionnaire developed by the study investigators (*Suppl Table 1*). The interviews were designed to gather information regarding (a) participants’ perceptions of common risks for stroke and high BP among AA adults aged 50 years and older living in their communities, (b) their self-management needs for HTN and post-stroke life, (c) preferred intervention components to support them in navigating the challenges of post-stroke life, (d) barriers to accessing healthcare services, and (e) strategies for future community-engaged interventions. Transcriptions were completed within 72 hours of each interview. As new themes emerged, the interview guide was refined to incorporate additional questions in subsequent interviews. Participants were compensated for their time by $50 gift cards at each data collection time point.

**Table 1:** Characteristics of the Participants with Stroke (N=25)

| **Participant Characteristics** | **n (%)** |
| --- | --- |
| **Age** |  |
| 50-65 | 10 (40) |
| 66-75 years | 12 (48) |
| 76 and above | 3 (12) |
| **Marital status** |  |
| Married | 12 (48) |
| Divorced/Widow/Single | 12 (48) |
| Missing | 1 (4) |
| **Education** |  |
| College level graduation | 8 (32) |
| High school graduation | 16 (64) |
| Missing | 1 (4) |
| **Income** |  |
| < 10K | 3 (12) |
| 10K – 30K | 11 (44) |
| 31K and above | 8 (32) |
| Missing | 3 (12) |
| **Living alone** |  |
| Yes | 15 (60) |
| No | 10 (40) |
| **Health Insurance** |  |
| Yes | 23 (92) |
| No | 1 (4) |
| **Self-rated Health** |  |
| Good to excellent | 8 (32) |
| Fair to poor | 17 (68) |
| **Years with HTN** |  |
| 3-5 | 3 (12) |
| 6-7 | 2 (8) |
| 8-10 | 3 (12) |
| Above 10 years | 15 (60) |
| **Years with Stroke** |  |
| 1-3 | 5 (20) |
| 3-5 | 8 (32) |
| 6-8 | 10 (40) |
| 8-10 | 2 (8) |
| **Location of residence** |  |
| Hale County | 7 (28) |
| Sumter County | 6 (24) |
| Pickens County | 12 (48) |

### Data Analysis

#### Quantitative analysis

Survey data were analyzed using univariate descriptive statistics using SPSS software (version 29.0). Frequencies and percentages were calculated to summarize participants’ sociodemographic characteristics. For the HK-LS, mean and standard deviation values were calculated for total scores (range 0-22). Participants’ knowledge levels were categorized as low (0-11) or high (>11). Knowledge levels for each subdimension were also classified as low or high, with scores below the midpoint indicating low knowledge.

Descriptive statistics were used to assess adherence levels based on scores from the *H-SCALE*. Participants were classified as adherent if they scored 21 on the medication subscale (range: 0– 21), >7 on the physical activity (PA) subscale (range: 0–14), and >40 on the weight management subscale (range: 10–50). Adherence to the DASH diet was categorized as high (51–77), moderate (33–50), or low (<32). Participants who reported no smoking or alcohol consumption received a score of zero on these subscales.

For the SSSMQ, participants’ self-management competency levels were classified as low (0–56), moderate (57–111), or high (>111) based on total scores. Frequencies and percentages were calculated for each category.

#### Qualitative analysis

Interview data were transcribed and analyzed using inductive thematic analysis, following a six-phase approach. In phase-1, senior research team members with expertise in qualitative analysis familiarized themselves with the interview transcripts and the interview notes made by the research assistants. In phase 2, senior research members independently generated initial codes from the data, followed by phase 3, where they then identified themes independently. During phase 4, researchers compared their codes and themes to ensure completeness and resolve discrepancies. Any disagreements were addressed in phase 5 with input from a third independent qualitative expert. In phase 6, final themes were reported and illustrated with direct participant quotes.

#### Triangulation and Integration of Results

To ensure rigor and validity of findings, qualitative and quantitative data were integrated using triangulation strategies. Findings from both qualitative and quantitative data were analyzed separately, then compared and synthesized to draw meaningful inferences. This approach facilitated a comprehensive understanding of participants’ experiences and strengthened the validity of the conclusions.

## Results

Table 1 displays social and demographic information about study participants.

Almost half (48%) of the participants were aged 66-75 years old and over three quarters (76%) lived alone. Three participants (12%) had an annual household income below $10K, while most (44%) had an income between $10K and $30K. Most participants (93%) reported having no college education. Additionally, 92% of participants had health insurance, and the majority (72%) rated their health as poor. Furthermore 60% of participants had been living with HTN for more than 10 years, and 40 % reported having had a stroke during the past 6 to 8 years.

Table 2 presents participants’ knowledge of HTN. The overall mean knowledge score was 29.05 (SD = 4.27), with the majority (76%) demonstrating low overall knowledge. Specifically, most participants had limited knowledge about the definition of HTN (76%) and its complications (64%). However, higher knowledge levels were observed in areas related to lifestyle modifications (78%), adherence to the Dietary Approaches to Stop Hypertension (DASH) diet (74%), and medication compliance (64%).

**Table 2:** Participants’ Knowledge Related to HTN (N=25)

| **Knowledge scores** | **Mean (S.D.)** |
| --- | --- |
|  | 29.05 (4.27) |
| **Knowledge Levels** | **n(%)** |
| Low level (0-22) | 19(76) |
| High Level (23 and above) | 6 (26) |

Table 3 summarizes participants’ adherence to HTN self-management activities, as measured by the SSSMQ. The majority exhibited low adherence to prescribed HTN medications (76%), a healthy diet (84%), recommended physical activity levels (56%), and weight management behaviors (68%). Additionally, 72% reported smoking, while 64% reported abstaining from alcohol consumption.

**Table 3:**
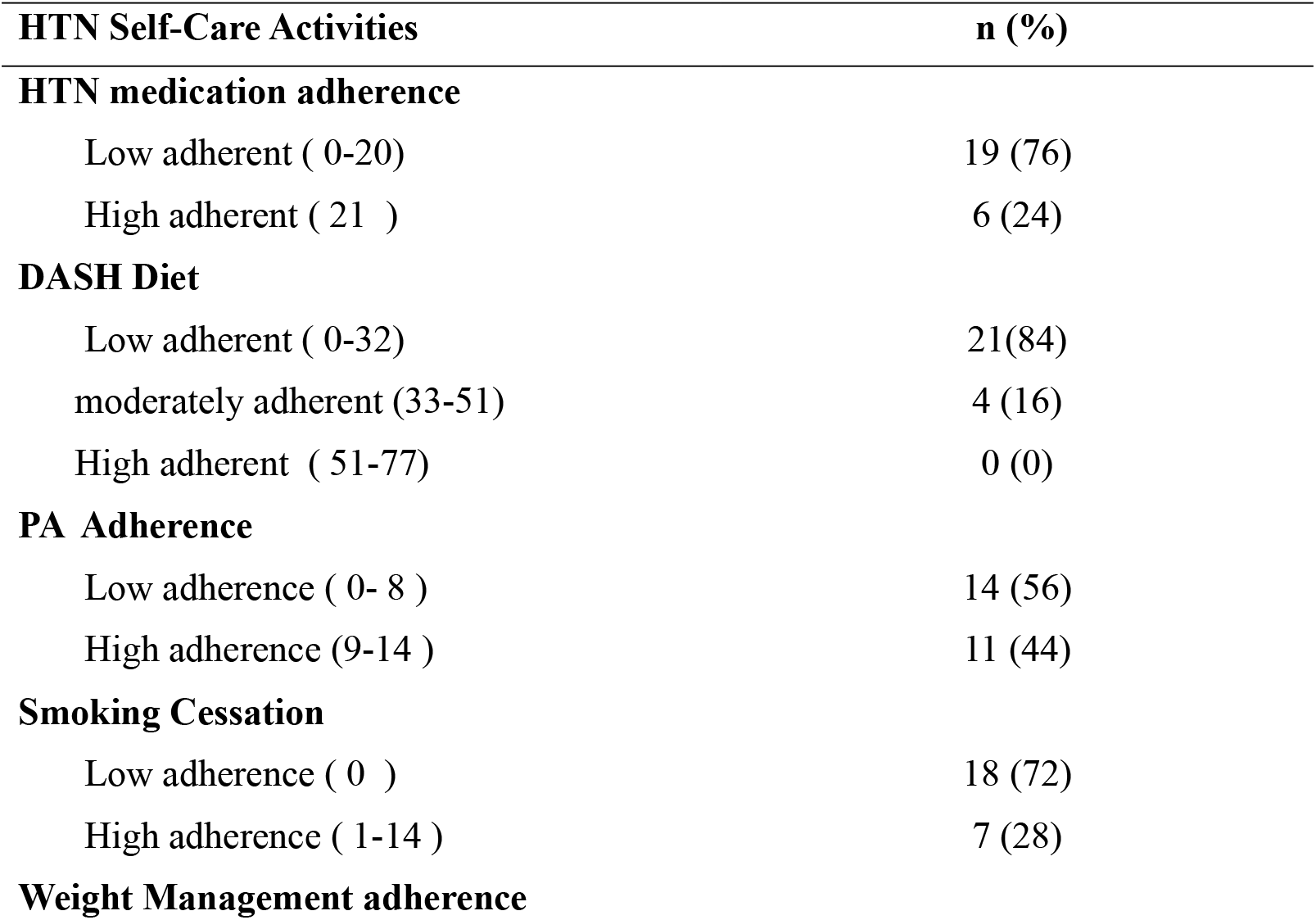
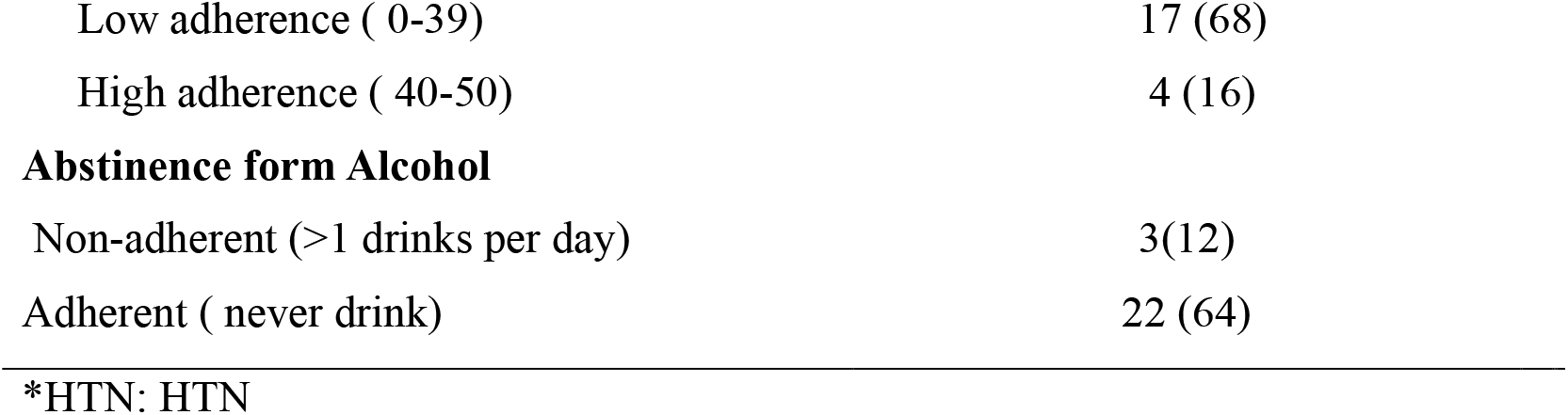
Participants’ Adherence Levels based on their HTN Self Care Activity Level Effect Scores (N=25)

| **HTN Self-Care Activities** | **n (%)** |
| --- | --- |
| **HTN medication adherence** |  |
| Low adherent ( 0-20) | 19 (76) |
| High adherent ( 21 ) | 6 (24) |
| **DASH Diet** |  |
| Low adherent ( 0-32) | 21(84) |
| moderately adherent (33-51) | 4 (16) |
| High adherent ( 51-77) | 0 (0) |
| **PA Adherence** |  |
| Low adherence ( 0- 8 ) | 14 (56) |
| High adherence (9-14 ) | 11 (44) |
| **Smoking Cessation** |  |
| Low adherence ( 0 ) | 18 (72) |
| High adherence ( 1-14 ) | 7 (28) |
| **Weight Management adherence** |  |
| Low adherence ( 0-39) | 17 (68) |
| High adherence ( 40-50) | 4 (16) |
| **Abstinence form Alcohol** |  |
| Non-adherent (>1 drinks per day) | 3(12) |
| Adherent ( never drink) | 22 (64) |

\*HTN: HTN

Table 4 describes participants’ overall competency in managing post-stroke life, ranging from 38 to 77, with a mean score of 59.0 (SD = 11.9). The majority (72%) demonstrated low competency levels, while 6 (24%) participants exhibited moderate competency. Only one participant achieved a high competency level for post-stroke self-management.

**Table 4:** Participants’ Stroke Self-Management Competencies to Manage Post-Stroke Life N=25.

| **Stroke Self-Management Competency** | **(n %)** |
| --- | --- |
| Low ( 0-56) | 18 (72) |
| Moderate (57-111) | 6 (24) |
| High (112-168) | 1 (4) |

### Qualitative Themes

#### 1. Lack of information/ health education

Participants indicated they had insufficient information to effectively manage their HTN after stroke. Most expressed confusion regarding stroke and its underlying mechanisms. They also reported a lack of knowledge about stroke symptoms, causes, and risk factors. Thus, a significant theme was a deficiency of information perceived by the participants. Many participants misunderstood the connection between stroke and HTN.

> “*After I had the stroke and came out of the hospital, maybe it would’ve been nice to have a doctor or a nurse to come in and- and just kind of check me and talk to me and bring some information. Some reading information that I could read over that would help me if-um, some other concerns came out about my health issues, but no-I didn’t-I didn’t get any follow-up or anything; no one check on me, no information was given*.*”*

They expressed concern about their knowledge gaps regarding medications, their side effects, and the importance of adhering to prescribed medication regimens. They also highlighted the lack of information about healthy lifestyle habits, including nutritious diet choices and physical activity. Some believed the resources available in their communities could be used to offer information and motivational programs for managing HTN and life after a stroke.

> *“Well, the provider I was going to-I wish that he had known to tell me that he was going to put me on* HTN *medication after my stroke. I would go to my appointments and each time he would say ‘your BP’s still up*.*’ And uh, they didn’t give me any meds, and they did not tell me what to do to control my BP, and what I could do to try to keep from having a stroke*.*”*

#### 2. Challenges associated with post-stroke life

Stroke survivors reported experiencing significant stress associated with life after a stroke. A major source of this stress was the changes in their roles, which they identified as a significant factor contributing to their difficulties. Financial burden also emerged as a common stressor, with participants associating their disabilities and reduced functional abilities to financial strain. While many survivor participants recognized the need for rehabilitation and recovery, most reported an inability to access these services due to lack of transportation and the financial burden of the associated costs and co-pays. This lack of access contributed to psychological stress as they were unable to pursue the care they needed. Some of the participants also mentioned the decrease in their quality-of-life post-stroke, reporting decreased enjoyment and socialization, while also experiencing increased stress as factors impacting their well-being.

“*I was prescribed with some medicine, but the co-pay for medicine was so expensive. I was not able to pay for it. It wasn’t doing no good to me*.*”*

“*Oh, man, emotionally you feel drained, uh, you feel picked on, you feel as though, a lot of times, you look at what and then how you have to struggle to do things that was really, really easy for you to do, you know, like I can’t even cut my grass anymore. I mean, watch your care things, you know, little things like it may not seem much, but I can’t do those things anymore. I told you it’s stressful, right? That’s a lot of strain on you. As I told you about my grandkids, you know, I just know if my health were better, I could get on the floor and play with them. I don’t know about nobody else, but you’d be a lot angrier when you can’t get things done quickly than you were before*.*”*

#### 3. Lack of continuity of care and communication with providers

Another common concern among rural stroke survivors was inadequate communication with their healthcare providers. Some participants reported feeling overlooked by providers when discussing their post-stroke challenges. Others mentioned experiencing delays in receiving timely treatment for their HTN. Participants expressed a need for rehabilitation services and continuity of care after discharge. Most believed their outcomes would have improved had they received adequate stroke rehabilitation, including physical therapy, occupational therapy, or speech therapy after their hospital discharge.

> “After having a stroke, *my blood pressure stayed elevated until I asked the question about the BP medicine again. I had to go through some channels to get the medicine approved, and after getting the medicine, my BP-the last time I went was the best it’s been in years*.*”*
>
> *“I wasn’t having therapy every day. It might be sometime twice a week or something like that, so that was it. They told me that’s all they offer. I felt like I needed more therapy; that is my struggle. I felt as though I needed supervised therapy; I just felt like if I could have had therapy constantly, every day, for five or six months, I could have done better*.*”*

Many participants recognized the need for follow-up and guidance from their healthcare providers to manage ongoing care needs after being discharged from the hospital following stroke treatment. However, they reported not receiving any follow-up to check on their treatment compliance or changes in their health status after discharge.

> “*The providers are not trying to sit down and tell you maybe you need to try this, or I’m going, give you a chance to try this medicine, they’re not doing that. And then, if you interrupt, they are going to make you feel like you are a fool or something. That’s why I say communication with patients is not good. I feel that when a doctor comes in and talks to you, they should sit there and listen to you because this is my body. If there’s something that could help me, you could tell me; sit down, talk to me, and tell me instead*.*”*
>
> *“They did not follow up after my discharge to home. I do go to the primary care provider, though. They did not teach me about the signs of stroke, what I should be careful of, and what to do for BP control, except my meds. Uh, the only thing I have was just access to the internet. That’s all I had. I get on there a lot of times now looking for stuff like walk-in tubs and any free equipment or anything that I could get to help me out. But you see different things that are online that could help, but all this stuff is so expensive”*.

#### 4. Lack of social support

Many of these individuals expressed concerns about their ability to perform daily activities after stroke. Some survivors reported a lack of support to perform these activities, while some participants indicated they had support from their family members. Most relied on a family member who lived separately to take them to healthcare provider appointments or refill their medications. Most stroke survivor participants received family support for transportation to various places.

Many participants reported feeling lonely as most lived away from their families and recognized the need to communicate with other community members, particularly fellow stroke survivors and their families. They noted the lack of a stroke support group in their rural areas. It was suggested that a communication system and a support group for stroke survivors and their caregivers be established.

> *“I think what would be most helpful to me would be the social groups so that, uh, you know, I can do face-on, uh, face-to-face conversation. That would be most helpful to me”*.

#### 5. Nonadherence to health behaviors

Participants expressed concern regarding their non-adherence to HTN self-management behaviors, such as physical activity, medication adherence, maintaining a healthy diet, and smoking cessation. They reported insufficient communication with their healthcare providers after their stroke, resulting in a lack of guidance for managing life challenges and treatment regimens following hospital discharge. They emphasized the need for rehabilitation and counseling services but noted the inaccessibility of these services, if available at all. Additionally, participants voiced concerns about the scarcity of stroke self-management programs in rural areas.

> *“My lifestyle and my behavior didn’t change, so even after I had a stroke, my eating habits are the same, and I am not exercising and stuff… and eventually, my weight picked up… and I gained just about 65 or 70 pounds. I just really didn’t take it seriously. Um, but it-scares me enough to try to try to do something, but I was a bad alcoholic too. So, a lot of my health issues result from my lifestyle*.*”*

## Discussion

The study found that most stroke survivors had low knowledge about hypertension (HTN) and its complications despite HTN being a major modifiable risk factor for stroke. This lack of awareness is common among survivors, especially in the “stroke belt,” which faces healthcare and socioeconomic challenges.^11, 12^ Without proper knowledge, individuals remain at higher risk for recurrent strokes.^13, 14^ Participants reported a significant decline in quality of life, with increased dependence on others and financial strain due to inability to work and high medication costs. A lack of understanding about stroke causes and prevention also highlighted gaps in education. These findings suggest the need for community-based education programs to address HTN knowledge, promote medication adherence and support survivors in managing physical, emotional, and financial challenges.

### Self-Care Behaviors for HTN Management

Effective HTN management relies on self-care behaviors such as medication adherence, weight management, dietary changes (e.g., DASH diet), physical activity, and reducing tobacco and alcohol use. Our survey revealed suboptimal adherence: 76% for medication, 84% for diet, 56% for PA, and 68% for weight management. Additionally, 72% smoked, and 36% consumed alcohol, underscoring HTN management challenges. The Southern dietary pattern, high in fried foods, processed meats, and sugary drinks, contributes to heart disease and obesity, particularly among AAs in the Stroke Belt. This diet is shaped by food insecurity, limited access to healthy foods, and financial constraints. ^15^

Increased physical activity reduces HTN and stroke risk. Still, leisure-time physical activity rates are lower in the Southern U.S. and among minority populations due to socioeconomic, environmental, and cultural factors, such as limited access to safe spaces and cultural attitudes toward exercise.^16^ Addressing these barriers through community-based advocacy, resource allocation, and health education is key to improving HTN self-care. ^17, 18^ Tobacco use^19^ and excessive alcohol consumption are major HTN risk factors,^20^ with tobacco use particularly prevalent in rural and older populations.^21^ Targeted interventions considering age and cultural context are needed.^22^ Additionally, inadequate patient education is a significant barrier to HTN management, particularly in underserved communities. Improving health literacy through educational interventions is crucial for better adherence and reducing HTN-related complications.

This study emphasizes the need for tailored interventions to improve HTN self-management. These interventions must address educational gaps, promote lifestyle changes, and leverage community resources for ongoing support, considering personal, social, and environmental factors. Addressing personal and environmental factors can foster greater self-efficacy and empower individuals to manage their HTN more effectively.

### Competencies for Self-Management of Post-Stroke Life

Post-stroke management is challenging, with survivors facing new roles, financial strain, and complex rehabilitation needs, leading to emotional distress and reduced quality of life. ^23 24^ The economic burden of medical care and rehabilitation often limits access to necessary services. In our study, most participants (72%) scored low in self-management, indicating they felt unprepared to manage their health post-stroke. Only 24% had moderate competency, and one participant showed high competency. As seen in other studies, these findings emphasize the need for targeted interventions to improve self-management skills and access to resources.^25^ Effective self-management programs should focus on skill-building, goal-setting, problem-solving, and developing coping strategies. Community-based resources offering home therapy, referral services, and health education can address key aspects of post-stroke care, such as skin breakdown prevention, endurance, adaptive equipment use, and daily living activities. ^4^

Evidence shows that community-based self-management programs enhance quality of life and self-efficacy, which improves occupational performance.^26^ A review by Khatri also found these programs address social determinants of health, promote well-being, reduce costs, prevent disease progression, and lower hospitalization rates. ^27^ The findings of this study underscore the need for comprehensive, community-based stroke programs that address not only the physical aspects of recovery but also the psychosocial, financial, and logistical challenges that stroke survivors face. Tailored interventions that integrate education, peer support, and practical resources—such as assistance with transportation and economic management—can enhance self-management competencies and overall quality of life. By addressing these multifaceted needs, such programs can better support survivors in navigating the complexities of life after stroke, leading to improved recovery outcomes and long-term well-being.

## Discussion of qualitative data themes and triangulation of findings

Participants indicated a paucity related to their knowledge of stroke, its risk factors, and the link between HTN and stroke. This finding was supported by our quantitative data indicating a low knowledge among 76% (19) of the participants. Our participant’s interview data indicated a lack of knowledge about lifestyle changes, which led to nonadherence to a healthy lifestyle like diet and physical activity. The survey data also showed that most participants had low adherence to a heart-healthy diet, 84% (21), physical activity, 56% (14), and weight management, 68% (17). Consequently, they reported minimal changes in lifestyle post-stroke. A recent systematic review reports similar findings of low knowledge among stroke survivors. The study also indicated the effect of educational intervention on knowledge, anxiety, and depression among these participants.^28^

Barriers to medication adherence, such as lack of co-pay, were common, with 76% of participants reporting low adherence. Participants also noted inappropriate medication doses, likely due to therapeutic inertia among primary care providers. Interview data highlighted challenges in transitioning care from medical facilities to home, including limited follow-ups, lack of home health care, and rehabilitation services. Other studies also report similar issues,^29 30 31^ with limited access to specialized stroke care and resources in rural areas hindering recovery. Addressing these barriers is crucial for promoting health in underserved rural populations.

Participants felt isolated in managing their risk factors, with loneliness and lack of social support hindering post-stroke life due to disabilities. About 60% of participants were living alone, and only half were married. Around 72% had low self-management competencies and needed support groups and home health visits. Other studies also highlight loneliness among rural stroke survivors,^32^ emphasizing the need for interventions to improve social support in these populations. ^30^

### Limitations and recommendations

This study has several limitations, including a small sample size; however, our findings were comparable to those in studies with larger samples. The study involves limitations of self-report data associated with this study; therefore, more objective data must be measured. There was a wide range of years between when participants had their stroke. A study with a more homogenous cohort can be conducted in the future.

## Conclusion

Community engagement, which encompasses social participation ^33^, community involvement ^34^, and access to supportive resources ^35^, is essential in the context of stroke survivors. It not only aids in recovery but also improves quality of life and fosters a sense of belonging among stroke survivors. Stroke survivors must be actively educated on the risk factors for stroke, possible reoccurrence, and how to manage the post-stroke life. Effective community engagement and support services are vital in assisting stroke survivors as they transition smoothly from hospital to home and reintegrate into their communities. Moreover, rural healthcare providers may have fewer resources and limited opportunities for continuing education on the latest stroke care protocols. Tackling these challenges necessitates innovative solutions, such as mobile health clinics, telehealth services, and community health worker programs, to bridge the gap in healthcare access and offer essential support to stroke survivors in rural areas.

## Data Availability

All data produced in the present study are available upon reasonable request to the authors

---

**Supplemental Table 1:**
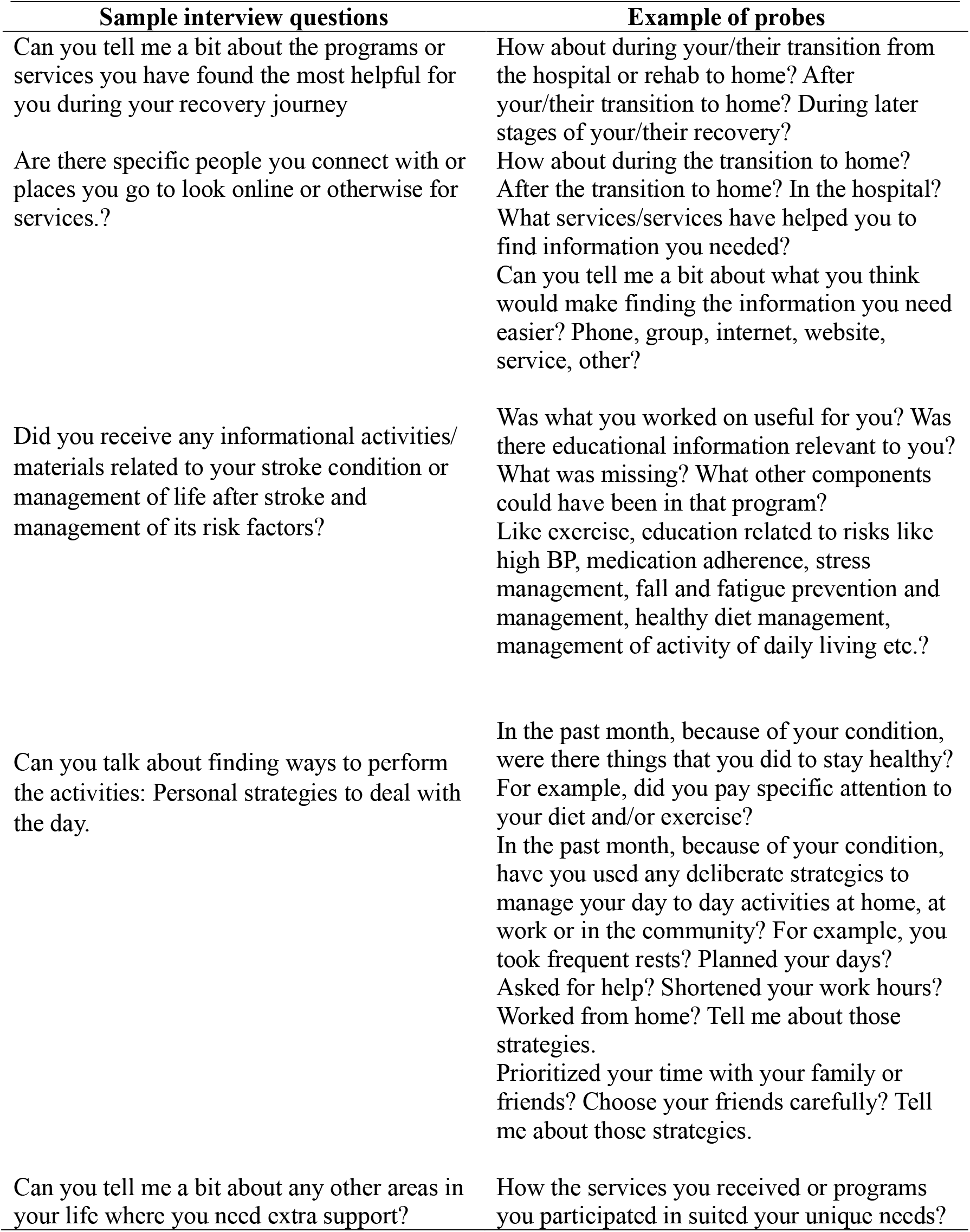
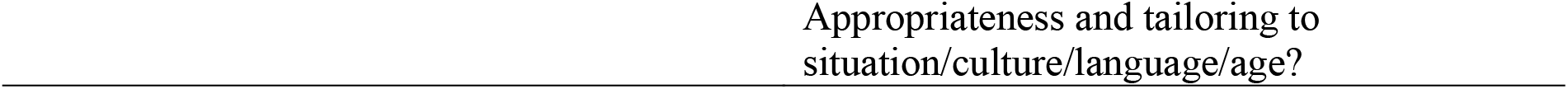
Semi–structured interview guide.

